# Machine learning to phenotype pain and predict response to pain interventions among young adults with irritable bowel syndrome

**DOI:** 10.1101/2025.10.07.25337516

**Authors:** Jie Chen, Aolan Li, Weizi Wu, Wanli Xu, Tingting Zhao, Angela R. Starkweather, Leonel Rodriguez, Ming-Hui Chen, Xiaomei S. Cong

## Abstract

**Introduction:** Irritable bowel syndrome (IBS) is a prevalent disorder whose most debilitating symptom is pain. The complex, multifactorial nature of IBS pain leads to highly variable and often inadequate responses to self-management, underscoring the urgent need for personalized prediction models.

**Methods:** This ancillary analysis of a randomized controlled trial (NCT03332537) utilized data from 80 young adults with IBS. We applied the Bayesian Additive Regression Trees machine learning algorithm to develop 27 distinct predictive models for pain severity, pain interference, and quality of life (QOL) at baseline and post-intervention. Predictors included a comprehensive, multi-domain set of variables spanning genetics, quantitative sensory testing, gut microbiota, psychosocial factors, and food intake.

**Results:** Model performance was strong, with area under the curve (AUC) values ranging from 0.753 to 0.981. A consistent hierarchy of predictors emerged. The *COMT* rs4680 polymorphism was the most significant predictor, featuring in 26 models, followed by *ADRA1D* rs1556832 in 24 models. The mechanical pain threshold was a key predictor of pain severity, while psychosocial factors, particularly pain catastrophizing, were crucial for pain interference and QOL. Gut microbiota features and food intake were also consistently important.

**Discussion:** This study establishes a comprehensive, multi-omics framework that explains individual differences in IBS pain and treatment response. The identified predictors provide a practical tool for advancing precision medicine. By classifying patients based on their distinct profiles, clinicians can proactively customize self-management strategies, potentially transforming care for this complex condition.

**WHAT IS KNOWN:**

- IBS pain involves complex, dysregulated gut-brain interactions.
- Treatment response is highly variable and difficult to predict.
- Machine learning can help phenotype complex pain conditions.

**WHAT IS NEW HERE:**

- Machine learning based hierarchical models identify a core neurogenetic predisposition for IBS pain.
- Machine learning based prediction of individual response to self-management before treatment begins.
- This provides a direct pathway to precision pain management in IBS.

## 1. Introduction

Irritable bowel syndrome (IBS) is a prevalent disorder of gut-brain interaction, affecting 8-12% of the global population.^1–4^ Its significant economic burden is matched by the profound impact of its cardinal symptom: recurrent abdominal pain. This pain is the primary reason patients seek specialist care and is consistently reported as the most distressing aspect of the condition, severely impairing quality of life. Various non-pharmacological interventions, including dietary changes, psychological therapies, and self-management strategies, are central to IBS pain management.^5–7^ These approaches leverage the therapeutic patient-provider relationship and empower patients with self-regulation skills.^8–12^ While studies, including our previous randomized controlled trial, confirm that such interventions can improve pain outcomes, patient responses are highly variable.^13,14^ The challenge lies in the complex, multifactorial nature of IBS pain. ^15,16^ Its pathobiology involves intricate, non-linear interactions between gut microbiota, host immune and nervous systems, diet, and psychosocial factors.^17–24^ Traditional statistical methods often fail to capture these complex relationships. Machine learning (ML) models, however, are ideally suited for this task. Algorithms like Bayesian Additive Regression Trees (BART) can integrate diverse data sources to identify nuanced patterns and interactions. This study leveraged data from a clinical trial (NCT03332537) to apply BART for phenotyping IBS pain and predicting treatment response by integrating a comprehensive set of variables, including sociodemographic, psychosocial, and genetic data, alongside pain sensitivity metrics and gut microbiota profiles.

## 2. Methods

Details of the design, rationale, and primary results of the RCT (NCT03332537) have been published.^12,25^ De-identified data were used in this analysis, including 1) demographic characteristics, 2) Brief Pain Inventory (BPI), 3) IBS quality of life (QOL), 4) Food Frequency Questionnaire (FFQ), 5) IBS-related psychosocial factors measured by (PROMIS) measures, Chronic Pain Self-Efficacy Scale (CPSES), and Coping Strategies Questionnaire-Revised (CSQ-R), 6) gut microbiota profile from stool samples, 7) quantitative sensory testing (QST), measured at baseline, and 6- and 12-week follow up; and pain related Single-Nucleotide Polymorphisms (SNPs) measured at baseline (Supplementary file 1).

All statistical analyses were performed using R version 4.4.3. Bayesian Additive Regression Trees (BART)^26^ were employed to investigate the relationship between predictors and the outcomes of pain severity, pain interference, and QOL. Separate models were fitted for each time point (0, 6, or 12 weeks) and three modeling approaches were employed. A concurrent prediction model was employed to predict the outcome at a given visit (0, 6, or 12 weeks) using dynamic predictors from that same visit in addition to pain-related SNPs. A forward prediction model estimates the absolute value of the outcome at a future visit using dynamic predictors from an earlier visit (e.g., predicting the 6-week outcome from baseline predictors, or the 12-week outcome from baseline or 6-week predictors). A change prediction model specifically predicts the change in outcome between two visits (e.g., the change in pain severity from baseline to 6 and 12 weeks, and from 6 to 12 weeks) using predictors from an earlier visit. The predictor variables included coping strategies, psychological factors (anxiety, depression, fatigue, positive affect, and sleep), nutrition and supplement intake, demographic characteristics (gender, race, ethnicity, education, marital status, employment, age, and caregiver type), QST, gut microbiota (genus and species), and SNPs. To enable classification modeling, outcome variables were dichotomized using clinically and empirically informed thresholds: BPI severity and BPI interference were set to 1 if the score was ≥2 and 0 otherwise; QOL was set to 1 if the score was ≥75 and 0 otherwise. For change outcomes, 1 indicated a positive change in value (increase from the earlier to the later visit), whereas 0 indicated no change or a negative change. The models did not explicitly model within-subject correlation across visits. Longitudinal information in the forward and change models was incorporated only through lagged predictors or difference scores. Model performance was assessed using the area under the receiver operating characteristic curve (AUC). The AUCs were calculated based on the posterior mean of BART-predicted probabilities. We applied 5-fold cross-validation across various BART hyperparameter settings (number of trees, node prior, and tree structure priors), selecting the configuration with the highest mean out-of-sample AUC. This configuration was then refit to the full dataset to obtain the final model. The top 20 influential predictors were identified based on their variable inclusion proportions, defined as the proportion of times a variable was used to split a decision node across all trees. The MCMC process included a burn-in period of 10,000 iterations, followed by posterior sampling with every 20th draw retained (thinning), yielding 10,000 posterior samples. Convergence was evaluated using trace plots and autocorrelation functions, confirming adequate mixing and stability of posterior draws.

## 3. Results

A total of 27 distinct predictive models for pain severity, pain interference, and QOL, both at baseline and for changes over time in response to interventions, were developed, including three concurrent prediction models (Figure 1a-c, Figure 2a-c, and Figure 3 a-c), three forward prediction models (Figure 1d-f, Figure 2d-f, and Figure 3 d-f), and three change prediction models (Figure 1g-i Figure 2h-i, and Figure 3g-i). The area under the receiver operating characteristic curve (AUC) for these 27 models ranged from 0.753 to 0.981 (Table 1), indicating good discriminative ability. The top 20 influential predictors of in the 27 models consistently included sociodemographic factors (such as self as the primary caregiver and/or education), psychological factors (coping strategies and self-reported psychological symptoms, e.g., depression and fatigue), measures of QST (e.g., mechanical pain threshold and cold pain threshold), SNPs (*COMT* rs4680, *ADRA1D* rs1556832, *TNFSF15* rs4263839), and specific genera and species of gut microbiota.

**Figure 1.**
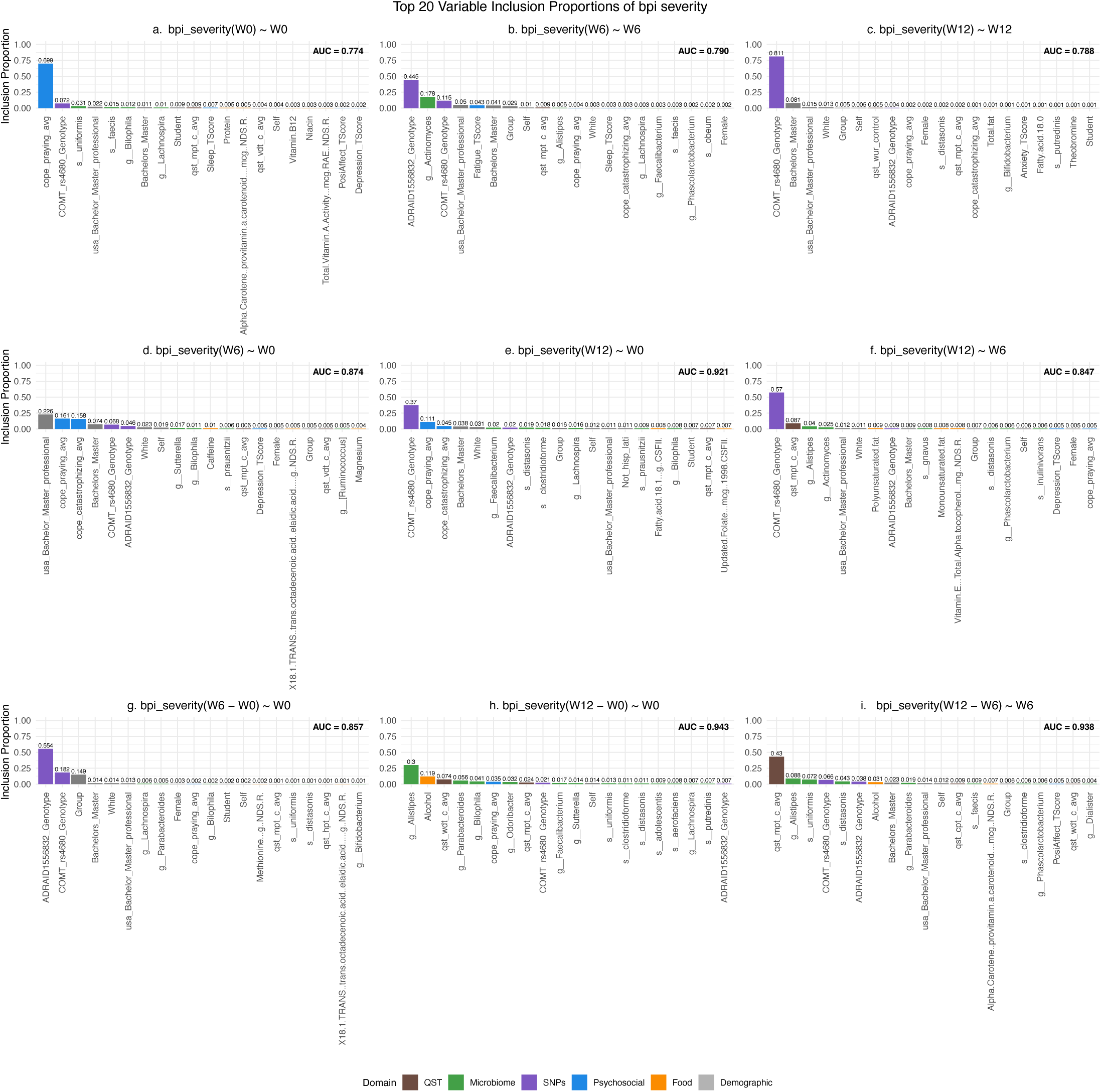
Biopsychological variables predict pain severity and changes to self-management interventions. 1a-c, concurrent prediction models were employed to predict the outcome at a given visit (0, 6, or 12 weeks) using dynamic predictors from that same visit in addition to pain-related SNPs. 1d-f, forward prediction models estimate the absolute value of the outcome at a future visit using dynamic predictors from an earlier visit (e.g., predicting the 6-week outcome from baseline predictors, or the 12-week outcome from baseline or 6-week predictors). 1g-i, change prediction models specifically predict the change in outcome between two visits (e.g., the change in pain severity from baseline to 6 and 12 weeks, and from 6 to 12 weeks) using predictors from an earlier visit.

**Figure 2.**
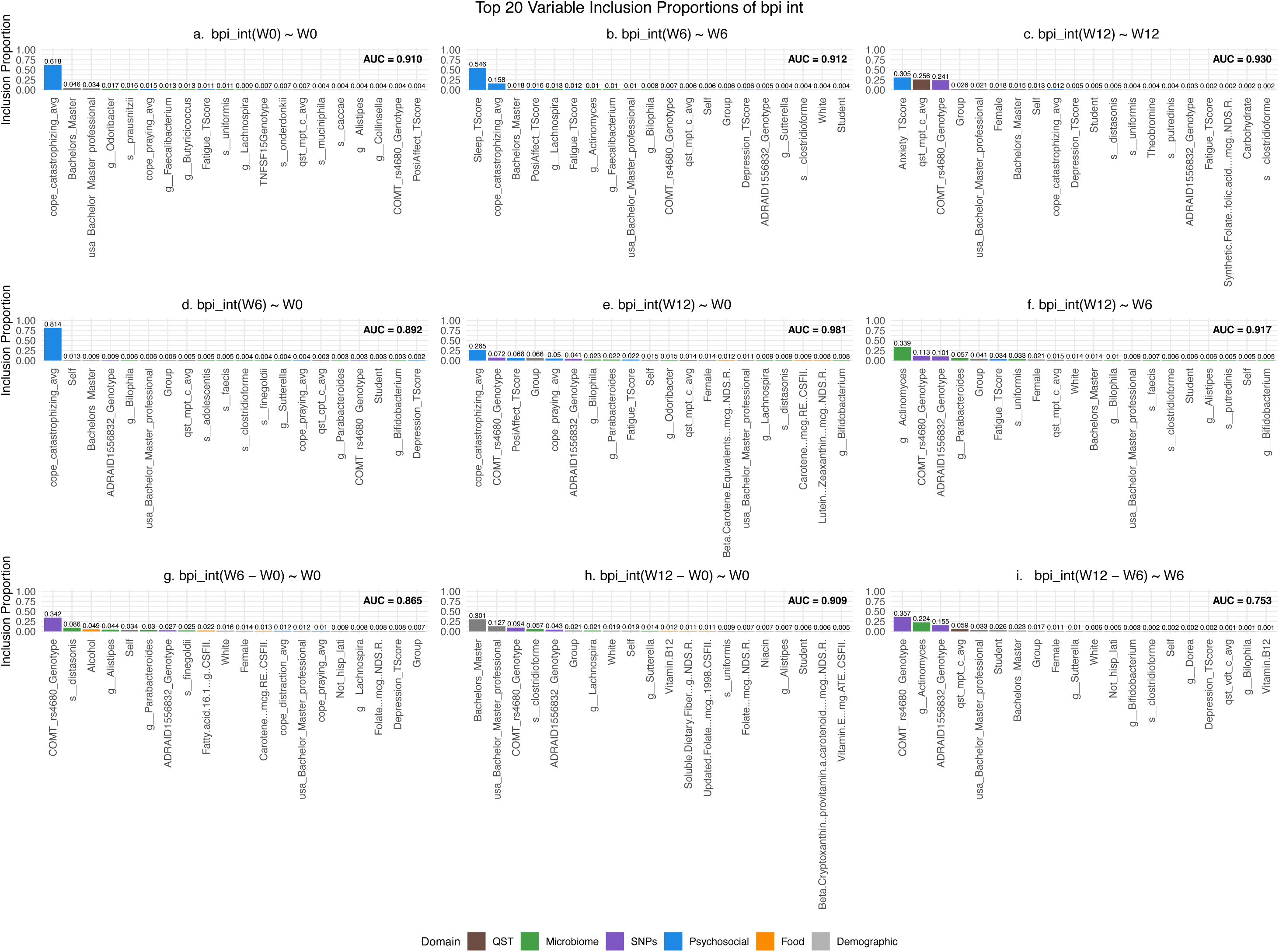
Biopsychological variables predict pain interference and changes to self-management interventions. 2a-c, concurrent prediction models were employed to predict the outcome at a given visit (0, 6, or 12 weeks) using dynamic predictors from that same visit in addition to pain-related SNPs. 2d-f, forward prediction models estimate the absolute value of the outcome at a future visit using dynamic predictors from an earlier visit (e.g., predicting the 6-week outcome from baseline predictors, or the 12-week outcome from baseline or 6-week predictors). 2g-i, change prediction models specifically forecast the change in outcome between two visits (e.g., the change in pain severity from baseline to 6 and 12 weeks, and from 6 to 12 weeks) using predictors from an earlier visit.

**Figure 3.**
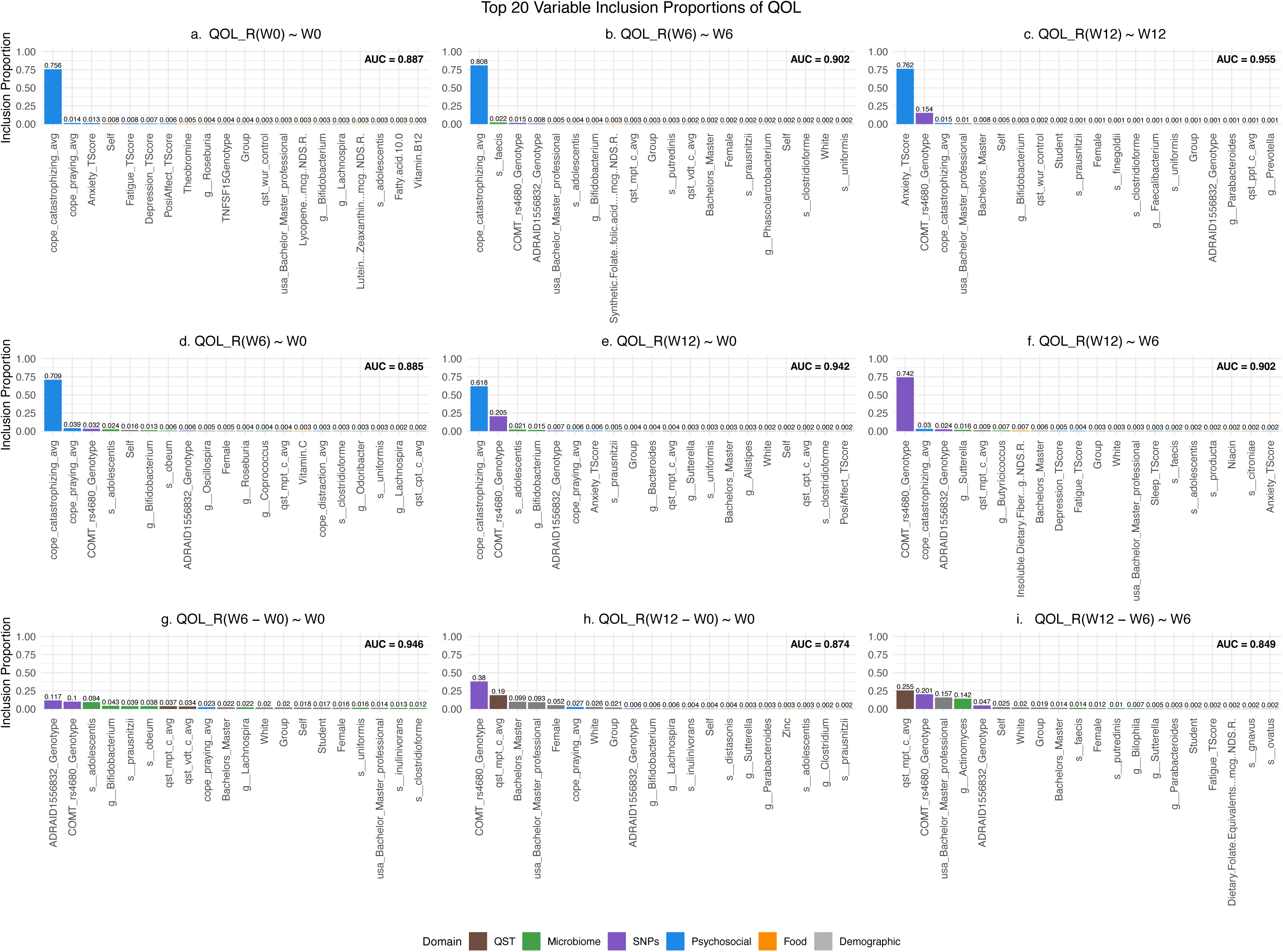
Biopsychological variables predict quality of life and changes to self-management interventions. 3a-c, concurrent prediction models were employed to predict the outcome at a given visit (0, 6, or 12 weeks) using dynamic predictors from that same visit in addition to pain-related SNPs. 3d-f, forward prediction models estimate the absolute value of the outcome at a future visit using dynamic predictors from an earlier visit (e.g., predicting the 6-week outcome from baseline predictors or the 12-week outcome from baseline or 6-week predictors). 3g-i, change prediction models specifically predict the change in outcome between two visits (e.g., the change in pain severity from baseline to 6 and 12 weeks, and from 6 to 12 weeks) using predictors from an earlier visit.

**Table 1.** The operating characteristic curve of the prediction models.

| Visit | AUC (in sample, out-of-sample) | AUC (in sample, out-of-sample) | AUC (in sample, out-of-sample) |
| --- | --- | --- | --- |
| **Pain severity** |  |  |  |
| Current prediction models <sup>a</sup> | **0\***<br>0.774 (0.828, 0.649) | **6\***<br>0.790 (0.784, 0.685) | **12\***<br>0.788 (0.792, 0.778) |
| Forward prediction models <sup>b</sup> | **0_6\***<br>0.874 (0.894, 0.784) | **0_12\***<br>0.921 (0.863, 0.709) | **6_12\***<br>0.847 (0.884, 0.674) |
| Change prediction models <sup>c</sup> | **0_6\***<br>0.857 (0.832, 0.644) | **0_12\***<br>0.943 (0.915, 0.665) | **6_12\***<br>0.938 (0.852, 0.634) |
| **Pain interference** |  |  |  |
| Current prediction models <sup>a</sup> | **0\***<br>0.910 (0.925, 0.814) | **6\***<br>0.912 (0.941, 0.789) | **12\***<br>0.903 (0.870, 0.732) |
| Forward prediction models <sup>b</sup> | **0_6\***<br>0.892 (0.917, 0.819) | **0_12\***<br>0.981 (0.978, 0.753) | **6_12\***<br>0.917 (0.912, 0.886) |
| Change prediction models <sup>c</sup> | **0_6\***<br>0.865 (0.830, 0.655) | **0_12\***<br>0.909 (0.914, 0.747) | **6_12\***<br>0.753 (0.731, 0.725) |
| **Quality of life** |  |  |  |
| Current prediction models <sup>a</sup> | **0\***<br>0.887 (0.898, 0.807) | **6\***<br>0.902 (0.900, 0.832) | **12\***<br>0.955 (0.937, 0.838) |
| Forward prediction models <sup>b</sup> | **0_6\***<br>0.885 (0.912, 0.791) | **0_12\***<br>0.942 (0.961, 0.904) | **6_12\***<br>0.902 (0.870, 0.739) |
| Change prediction models <sup>c</sup> | **0_6\***<br>0.946 (0.887, 0.781) | **0_12\***<br>0.874 (0.817, 0.700) | **6_12\***<br>0.849 (0.766, 0.716) |

Notes: 5-fold cross-validation was used to select the best hyperparameter configuration for each outcome-task combination (row) based on the mean out-of-sample operating characteristic curve (AUC) across visits (columns). The selected configuration was then applied to the whole dataset to obtain the final AUC (full AUC). Each cell shows [full AUC] with mean in-sample and out-of-sample AUCs in parentheses.
<sup>a</sup> Concurrent prediction models were employed to forecast the outcome at a specific visit (0, 6, or 12 weeks) using dynamic predictors from that same visit, along with pain-related SNPs.
<sup>b</sup> Forward prediction models estimate the actual value of an outcome at a future visit by using dynamic predictors from an earlier visit (e.g., predicting the 6-week outcome from baseline predictors or the 12-week outcome from baseline or 6-week predictors).
<sup>c</sup> Change prediction models specifically forecast the change in outcomes between two visits (e.g., the change in pain severity from baseline to 6 and 12 weeks, and from 6 to 12 weeks) using predictors from an earlier visit.
\*Data were collected at 0, 6, and 12 weeks.

### 3.1 Biopsychological variables predict pain severity and response to self-management interventions

*COMT* rs4680 was included among the 20 influential predictors in all nine models and had a proportion greater than 0.05 in eight models, except for the model that used baseline characteristics to predict changes in pain severity from baseline to 12 weeks (Figure 1h). *ADRA1D* rs1556832 was included in the 20 influential predictors in seven of the nine models and had a proportion greater than 0.05 in only one model, where it was the most significant predictor of changes in pain severity from baseline to 6 weeks (Figure 1g). The mechanical pain threshold (MPT) was among the 20 influential predictors in all nine models and was also the most significant predictor in the model predicting changes from 6 weeks to 12 weeks, using variables measured at the 6-week visit (Figure 1i). Food intake (dietary patterns) was among the top 20 predictors in 8 of the 9 models, except for the concurrent prediction model that estimated pain severity at 6 weeks based on variables measured during the 6-week visit and pain-related SNPs (Figure 1b).

Coping (praying) was included among the 20 most influential predictors in 8 of the nine models, except for the model that used characteristics measured at 6 weeks to predict changes in pain severity from 6 to 12 weeks (Figure 1i). Coping (catastrophizing) was only included among the top 20 influential predictors in four models, which utilized concurrent characteristics to predict pain severity at 6 and 12 weeks (Figure 1b and 1c) and baseline characteristics to predict pain severity at 6 and 12 weeks (Figure 1d and 1e).

The genus of gut microbiota included in the 20 influential predictors varied across models, with *Lachnospira* and *Bilophila* included in five models, *Alistipes* in four, and *Bifidobacterium* and *Sutterella* in two. Additionally, *Alistipes* was the most significant predictor in the model predicting changes in pain severity from baseline to 12 weeks, based on baseline characteristics and pain-related SNPs (Figure 1h).

### 3.2 Biopsychological variables predict pain interference and response to self-management interventions

*COMT* rs4680 was among the 20 most influential predictors in all nine models and had a proportion exceeding 0.05 in six of them (Figure 2c, 2d, 2f, 2g, 2h, and 2i). Additionally, *COMT* rs4680 was the most significant predictor in the model predicting changes in pain interference from baseline to 6 weeks, using all baseline characteristics (Figure 2g), and the change from 6 to 12 weeks, using 6-week measures and pain-related SNPs (Figure 2i). *ADRA1D* rs1556832 was included in 8 models but was not part of the model predicting baseline pain interference using baseline measures and pain-related SNPs (Figure 1a). *TNFSF15* rs4263839 was only included in the model predicting baseline pain interference with baseline measures and genetic profiles, with a proportion of 0.007 (Figure 2a). The MPT was also among the 20 influential predictors in seven of the nine models, except for the change prediction models that estimated the change of pain interference from baseline to 6 weeks and from baseline to 12 weeks using baseline measures and pain-related SNPs (Figure 2g and 2h), but had a proportion greater than 0.05 only in two models (Figure 2c and 2i).

Coping (catastrophizing) was among the 20 key predictors in five of the nine models and was the most significant predictor for pain interference at baseline, 6-week, and 12-week visits, based on baseline measures and pain-related SNPs (Figure 2a, 2d, and 2e). Coping (praying) also appeared in the 20 key predictors in five of the nine models and had a proportion of 0.05 in one model (Figure 2e). Notably, sleep (disturbance) and anxiety emerged as the most significant predictors for pain interference at the 6-week and 12-week visits, respectively, using concurrent measures and pain-related SNPs (Figure 2b and 2c).

Although the gut microbiota genus and species were included in all nine models, the genera and species in each model varied. Food intakes were included in five of the nine models (Figure 2c, 2e, 2g, 2h, and 2i); the specific components differed, and none had a proportion equal to or greater than 0.05. More importantly, group or interventional differences in pain interference were detected in 8 of the nine models, except in one model that predicted baseline pain interference using baseline measures and genetic profiles (Figure 2a).

### 3.3 Biopsychological variables predict quality of life and changes to self-management interventions

*COMT* rs4680 was included among the 20 influential predictors in 8 of the 9 models and had a proportion greater than 0.05 in six models, but not in the model predicting baseline quality of life using baseline characteristics and pain-related SNPs (Figure 3a). Additionally, *COMT* rs4680 was the most significant predictor in two models: one predicting quality of life at 12 weeks using 6-week characteristics and pain-related SNPs (Figure 3f), and another predicting changes in quality of life from baseline to 12 weeks based on baseline characteristics and pain-related SNPs (Figure 3h). *ADRA1D* rs1556832 was also included in 8 models and was the most significant predictor in the model predicting changes in quality of life from baseline to 6 weeks using baseline characteristics and pain-related SNPs (Figure 3g). *TNFSF15* rs4263839 was only included in the model predicting baseline quality of life with baseline characteristics and pain-related SNPs, with a proportion of 0.004 (Figure 3a). The MPT was included among the 20 most influential predictors in 7 of the 9 models and was the most significant predictor in the model forecasting changes in quality of life from the 6-week visit to the 12-week visit, using measures from the 6-week visit and pain-related SNPs (Figure 3i).

Coping (catastrophizing) was included among the 20 most influential predictors in 6 of the 9 models and was the top predictor in models forecasting quality of life at baseline, 6-week, and 12-week visits using baseline characteristics and pain-related SNPs (Figure 3a, 3d, and 3e). Additionally, Coping (catastrophizing) was also the strongest predictor in models predicting quality of life at 6 weeks using characteristics measured at the 6-week visit and pain-related SNPs (Figure 3b). Similarly, coping (praying) was included among the 20 influential predictors in 5 of the nine models, with none having a proportion equal to or greater than 0.05. Notably, anxiety was the most significant predictor in the model predicting quality of life at 12-week visits by using measures at the 12-week visit and pain-related SNPs (Figure 3c).

Although the gut microbiota genus and species were included in all nine models, the genus and species varied in each model. Food intakes were included in 6 of the nine models, with different components in each, and none had a proportion equal to or greater than 0.05 (Figure 3a, 3b, 3d, 3f, 3h, and 3i). More importantly, differences in group or interventional quality of life were detected in 8 of the nine models, except for those predicting quality of life at the 6-week visit using baseline characteristics and pain-related SNPs (Figure 3d).

## 4. Discussion

To our knowledge, this study is among the first to leverage machine learning for integrating diverse data types—demographics, diet, psychosocial factors, genetics, gut microbiota, and QST—to phenotype pain and QOL in young adults with IBS. By modeling both baseline states and responses to nurse-led pain self-management interventions, we have developed precise phenotypic profiles that offer deeper insights into the condition’s underlying mechanisms. Importantly, our models show that psychological factors (coping strategies, depression, and anxiety), genetic factors (*COMT* rs4680, *ADRA1D* rs1556832), QST profiles (such as MPT), and gut microbiota (like *Alistipes*) are not only baseline predictors but may also influence intervention outcomes. This extends beyond the traditional view that intervention effects are primarily psychosocial, suggesting a more complex biopsychosocial model of response.

This work builds upon prior evidence of the biopsychosocial complexity of IBS pain.^22,27–29^ By employing a comprehensive machine-learning framework across 27 predictive models, we move from describing complexity to delineating a hierarchical and predictive structure of IBS pain. Our findings integrate and extend previous observations into a coherent model where a core neurogenetic predisposition is modulated by cognitive-affective processes and gut-brain axis inputs.

The most striking discovery is the central role of catecholaminergic pathways. The *COMT rs4680* (Val158Met) polymorphism consistently appeared across models, underscoring its role as a key regulator of IBS pain. COMT influences the metabolism of catecholamines; the Met allele reduces enzymatic efficiency, elevating synaptic norepinephrine and dopamine and predisposing individuals to central sensitization.^30,31^ This aligns with recent meta-analytic evidence linking the COMT Met allele to heightened pain sensitivity across conditions.^32^ This further shaped by ADRA1D rs1556832, which encodes the alpha-1D adrenergic receptor,^33^ a main target of norepinephrine. Together, COMT and ADRA1D variants represent a “dual-hit” mechanism, amplifying adrenergic signaling and driving pain severity in a subset of IBS patients.^34^ The inclusion of ADRA1D in nearly all models supports adrenergic signaling as a fundamental mechanism, suggesting that targeted alpha-1 blockade could be a viable therapeutic strategy.^35^

This genetic risk translates into measurable physiology. The mechanical pain threshold (MPT) emerged as a strong predictor of pain severity, functionally linking genetic variants to central sensitization. Individuals with high-risk genetic profiles showed increased mechanical pain sensitivity on the non-dominant forearm, indicating body-wide hyperalgesia. The consistent presence of MPT across all nine pain-severity models validates that genetic predispositions (COMT and ADRA1D) manifest as systemic differences in pain processing. These results highlight a neurobiological basis for symptom variability and suggest the need for stratified treatment approaches targeting endogenous pain control pathways.^17,18^

Beyond the genetic-physiological axis, our models clarify how psychological processes shape the pain experience. Coping strategies, particularly catastrophizing, repeatedly appeared as influential predictors, reinforcing evidence that these cognitive factors strongly mediate pain reduction.^12,36,37^ Their inclusion alongside MPT in models for pain interference and quality of life illustrates a mechanistic flow: genetic and physiological factors establish baseline sensitivity, while psychological processes determine the lived impact of pain. Gut microbiota and diet also emerged as consistent predictors, emphasizing the gut-brain axis as a dynamic modulator.^29,38^ This integration of biological, psychological, and environmental influences suggests that lifestyle and microbial factors continually interact with an individual’s innate genetic and psychological profiles to shape clinical outcomes.

A key clinical breakthrough lies in predicting treatment response. The ability of multi-omics profiles to forecast changes following self-management interventions provides a mechanistic explanation for our prior RCT findings. It shows that patients with a high-risk profile—defined by the *COMT* Met allele, *ADRA1D* risk variant, low MPT, and high catastrophizing—are a subgroup for whom standard self-management may be ineffective unless it effectively addresses these specific biological and psychological factors.

The specific gut microbial taxa identified varied across models, indicating complex and individualized interactions that require further investigation. Future research should aim to identify the core functional pathways shared by these different microbial signatures. Our recent findings, using a subset of the dataset, also showed that a nurse-led self-management intervention improves IBS symptoms by affecting specific host-transcriptomic and gut-microbial pathways, even with a small sample size.^24^ Based on results from the pilot study, which only included participants with daily internet access, irritable bowel syndrome subtypes (e.g., constipation-predominant, diarrhea-predominant, mixed) were not considered as predictors in the current analysis. Future translational studies could also confirm these predictive insights within a stratified care model, testing whether assigning mechanism-specific treatments, such as adrenergic blockers, cognitive-behavioral therapy, or an intensive self-management program for high-risk patients (*COMT* Met allele, *ADRA1D* risk variant, increased mechanical pain threshold, and high catastrophizing) results in better outcomes compared to standard care.

## 5. Conclusion

In summary, this study creates a unified, hierarchical model of IBS pain by combining multi-omics data within a machine learning framework. By developing predictive models for pain trajectories and treatment responses using extensive longitudinal data from a randomized controlled trial, we identify key predictors—including *COMT* rs4680, *ADRA1D* rs1556832, mechanical pain threshold, and pain catastrophizing—that account for individual differences in self-management outcomes. These findings offer a mechanistic understanding of biopsychosocial interactions in IBS and serve as a practical tool for advancing precision medicine. The models developed here have direct translational potential, allowing clinicians to identify patients at high risk for poor outcomes and tailor personalized management strategies from the outset, ultimately moving IBS management beyond a one-size-fits-all approach.

## Author Contributions

Conceptualization, J.C., M.H.C., and X.C.; formal analysis, J.C., A.L., W.W., Z.T., M.H.C., and X.C.; funding acquisition, X.C., and A.S.; methodology, J.C., A.L., W.W., Z.T., W.X., A.S., M.H.C., and X.C.; project administration, J.C., W.X., A.S., and X.C.; writing-original draft, J.C., A.L., and X.C.; writing-review and editing, All. All authors have reviewed the manuscript and agreed to submit this version.

## Acknowledgments

The authors would like to acknowledge all the participants in this study. The authors would also like to acknowledge the support from the Bio-Behavioral Lab (BBL), the Center of Advancement in Managing Pain (CAMP), and the NIH-funded P20 Center for Accelerating Precision Pain Self-Management in the University of Connecticut School of Nursing. The authors would like to acknowledge the support from the Microbial Analysis, Resources, and Services (MARS) and the Institute for Systems Genomics (ISG) at the University of Connecticut.

## Conflicts of Interest

The authors declare no conflict of interest.

## Funding

This study was supported by the National Institute of Nursing Research of the National Institutes of Health (NIH-NINR) under award number: NIH-NINR P20NR016605 (PI Starkweather; Pilot PI: Cong) and NIH-NINR R01NR016928 (PI: Cong). Jie Chen received research support from the Florida State University First Year Assistant Professor (FYAP) Program. The funding agencies have no role in the design, conduct, or analysis of this study.

## Data Availability Statement

The datasets generated and analyzed in the current study are available from the NCBI dBGaP (https://submit.ncbi.nlm.nih.gov/subs/sra/SUB8914789/). Deidentified data will be available upon reasonable request. Requests to access these datasets should be directed to.

## Supplementary file 1

### Methods

#### 1. Study Design and Participants

Details of the design, rationale, and primary results of the RCT (NCT03332537) have been published.^1,2^ The RCT protocol was approved by the University Institute Review Board (IRB). Informed consent was acquired from each participant. Privacy was maintained during the entire data collection and management process. A total of 80 young adults with IBS were recruited in the northeastern region of the United States in this RCT. Young adults were enrolled in the study if they were (1)18-29 years of age; (2) able to read and speak in English; (3) able to access the internet; (4) having a clinical diagnosis of IBS according to the Rome III or IV criteria from a healthcare provider (for IBS subjects only). Subjects were excluded if they had (1) chronic pain conditions (e.g., chronic pelvic pain, headache, back pain, etc.); (2) severe mental health conditions; (3) celiac disease or inflammatory bowel disease; (4) infectious diseases; (5) diabetes mellitus; (6) injury or open skin lesions on the non-dominant arm; (7) history of substance abuse or regular opioid use; (8) history of prebiotics/probiotics or antibiotics use in the past months.^2^ Women during pregnancy or within 3 months postpartum period were also excluded. Allocation of the eligible participants with IBS to each intervention group (Online Modules vs. Nurse-Led Online Modules) was completed using a stratified and blocked randomization scheme. The full description of randomization and blinding was previously described in the published study protocol.^1^ De-identified data were used in this analysis.

#### 2. Assessment of Variables

This analysis leveraged data collected in the RCT (NCT03332537). Measurements in the RCT (NCT03332537) included demographic characteristics, pain, quality of life, symptoms, self-efficacy, and coping. The demographic characteristics include age, sex, ethnicity, race, education level, and other factors that were measured by the National Institute of Nursing Research common data elements.^3^ Questionnaires were completed online through the REDCap system using a laptop or iPad. The participants with IBS completed all questionnaires at enrollment (T0, 0 weeks), as well as at 6- and 12-week follow-up visits (T1 and T2, respectively).

##### Pain and self-reported symptoms

The average pain severity and pain interference were measured using the Brief Pain Inventory (BPI). The BPI has questions with 0-10 rating scales, where a higher score refers to more severe pain.^4^

##### Quality of life

IBS quality of life was measured by a 34-item IBS-specific quality of life instrument.^5^ This instrument was designed to capture patients’ perceptions of their daily functions interfered with by IBS. There are 8 subscales in this five-point Linkert instrument. The score range of this scale is 0-100, which higher score refers to a better quality of life.

##### Food Frequency Questionnaire (FFQ)

The self-reported FFQ was used to measure the type and quantity of food intake.^6^ The reliability of the FFQ was established by the National Health and Nutrition Examination Survey (NHANES II).^7^ In the Women’s Health Initiative study, the test-retest reliability of the nutrient intake estimates from the FFQ was reported as high with intra-class correlation coefficients ranging from 0.67 to 0.92.^8^ A higher number in each of the measures means a higher intake of the nutrition elements.

##### IBS-related psychosocial factors

IBS-related symptoms, including anxiety, depression, fatigue, and sleep disturbance, were measured by using the NIH Patient-Reported Outcomes Measurement Information System (PROMIS®) and scored following the system instruction.^9^ A higher T score PROMIS measurement indicated a higher intensity of the measured symptom, and a mean score greater than 55 indicates that a study subject experienced a significantly higher intensity of the symptom than those of the healthy reference population according to the PROMIS guide. Self-efficacy was measured by the 6-item Self-Efficacy for Managing Chronic Disease (SEMCD).^10^ The score of the Likert SEMCD ranged from 0 to 10. A higher score of SEMCD indicates great self-efficacy. Coping strategies were evaluated by the Coping Strategies Questionnaire-Revised (CSQ-R) to assess six cognitive coping strategies to pain, including distraction, catastrophizing, ignoring pain sensations, distancing from pain, coping self-statements, and praying.^11^ Each subscale has a score range of 0-36, with a higher score indicating a better coping strategy.

##### Buccal swab samples collection and pain related Single-Nucleotide Polymorphisms (SNPs) assay

Genotyping of selected pain-related SNPs was performed using buccal swab samples. Participants rinsed their mouths twice with water and then collected buccal cells by firmly brushing the inside of the cheek. The buccal samples were stored at −80 °C in a Biobehavioral Laboratory until further analysis. Genomic DNA was extracted from these samples using the Gentra Puregene Buccal Cell Kit, according to the manufacturer’s protocol (#158845).^12^ Eleven candidate polymorphisms were assessed, including *ADRA1D* rs1556832, *TNFSF15* rs4263839, *COMT* (rs4680, rs4818, rs6269, and rs4633), *5-HTTLPR* in *SLC6A4*, *HTR3A* rs1062613, *OPRM1* rs1799971, and *OXTR* (rs53576 and rs2254298). ^1,12^ Taqman SNP genotyping assays (VIC/FAM) and allelic discrimination were conducted on a StepOnePlus^TM^ PCR system using StepOne^TM^/StepOnePlus^TM^software v2.0 (Thermo Fisher Scientific, Waltham, MA, USA).

##### Stool sample collection and gut microbiota data sequencing

Stool samples were collected at home by the participants following our previous protocol using the OMNIgene®-gut (DNA Genotek, Ottowa, Canada) kit.^13,14^ Training on collecting the samples at home was provided to the participants by research personnel. After collection, participants were instructed to mail the samples back to the research laboratory within a week of the baseline visit data collection using a pre-labeled and pre-paid envelope with a tracking barcode. Once received by the laboratory, the stool samples were aliquoted into small tubes and stored in a −80 ℃ freezer for bulk processing. A total of 0.25 g stool from each participant was placed into a beat tube and sent out for DNA extraction and sequencing. The 16S rRNA V4 region amplicon was sequenced by the Illumina Miseq 2000 platform (Illumina, San Diego, CA) at the University of Connecticut (UConn) Microbial Analysis, Resource, and Services laboratory following our previous protocol.^14^ The raw 16S rRNA sequencing data were processed by the Mothur 1.43.0 software following the analysis pipeline of Miseq (http://www.mothur.org/wiki/MiSeq_SOP) to obtain the taxonomy and diversity of the gut microbiota. Paired-end sequences were combined into contigs and aligned against the SILVA 132 V4 16 S rRNA gene reference alignment database with poor-quality sequences removed. Operational taxonomic units (OTUs) were determined at a 97% identity.^15,16^

**Quantitative sensory testing (QST)** included 13 parameters, which were performed in the original study according to the published protocol.^28^ All QST tests were conducted on the skin of the subject’s upper limbs. A standard set of round-tipped von Frey-like monofilaments, 0.5 mm in diameter (Optihair2-Set, Marstock Nervtest, Germany), exerted standardized forces ranging from 0.25 to 512 millinewtons (mN) against the participant’s skin to assess mechanical pain sensitivities, including mechanical detection threshold (MDT), mechanical pain threshold (MPT), mechanical pain sensitivity (MPS), and wind-up ratio (WUR). A standardized brush (cleaned with antiseptic toilette between tests) was used to evaluate dynamic mechanical allodynia (ALL) by applying three light tactile stimuli to the subject’s medial forearm skin. A Rydel-Seiffer tuning fork (64 Hz, 8/8 scale) measured vibration detection threshold (VDT). The Medoc System™ (Medoc Ltd., Ramat Yishai, Israel) was utilized to determine thermal pain thresholds, such as cold detection threshold (CDT), warm detection threshold (WDT), cold pain threshold (CPT), and heat pain threshold (HPT). The pressure pain threshold (PPT) was assessed with a Medoc algometer (contact area 1.7 cm²), with a baseline pressure of 50 kPa, an average ramped rate of 30 kPa/second, and a cutoff of 600 kPa. The interval between each trial was 5-10 seconds. For each indicator, the results were averaged over three consecutive measurements taken on three different sites on the non-dominant forearm.

#### 3. Data analysis

The final analytical sample consisted of 80 participants. The cohort was predominantly female (n=61, 76.3%), non-Hispanic/Latino White, and comprised students who had received a college or associate degree; most participants identified themselves as their own primary caregivers. Participants were randomized to either an Online Modules group (n=41) or a Nurse-Led Online Modules group (n=39). All 80 participants contributed baseline (T0) data. At the 6-week follow-up, 35 participants (85.4%) in the Online Modules group and 27 (69.2%) in the Nurse-Led group provided data. At the 12-week endpoint, 30 (73.2%) and 26 (66.7%) participants completed data collection in the respective groups, yielding a total sample of 56.

All statistical analyses were performed using R version 4.4.3. Bayesian Additive Regression Trees (BART)^17^ were employed to investigate the relationship between predictors and the outcomes of pain severity, pain interference, and quality of life (QOL). Separate models were fitted for each time point (0, 6, or 12 weeks) and three modeling approaches were employed. A concurrent prediction model was employed to predict the outcome at a given visit (0, 6, or 12 weeks) using dynamic predictors from that same visit in addition to pain-related SNPs. A forward prediction model estimates the absolute value of the outcome at a future visit using dynamic predictors from an earlier visit (e.g., predicting the 6-week outcome from baseline predictors, or the 12-week outcome from baseline or 6-week predictors). A change prediction model specifically predicts the change in outcome between two visits (e.g., the change in pain severity from baseline to 6 and 12 weeks, and from 6 to 12 weeks) using predictors from an earlier visit. The predictor variables included coping strategies, psychological factors (anxiety, depression, fatigue, positive affect, and sleep), nutrition and supplement intake, demographic characteristics (gender, race, ethnicity, education, marital status, employment, age, and caregiver type), QST, microbiome (genus and species), and SNPs. To enable classification modeling, outcome variables were dichotomized using clinically and empirically informed thresholds: BPI severity and BPI interference were set to 1 if the score was ≥2 and 0 otherwise; QOL was set to 1 if the score was ≥75 and 0 otherwise. For change outcomes, 1 indicated a positive change in value (increase from the earlier to the later visit), whereas 0 indicated no change or a negative change. The models did not explicitly model within-subject correlation across visits. Longitudinal information in the forward and change models was incorporated only through lagged predictors or difference scores. Model performance was assessed using the area under the receiver operating characteristic curve (AUC). The AUCs were calculated based on the posterior mean of BART-predicted probabilities. We applied 5-fold cross-validation across various BART hyperparameter settings (number of trees, node prior, and tree structure priors), selecting the configuration with the highest mean out-of-sample AUC. This configuration was then refit to the full dataset to obtain the final model. The top 20 influential predictors were identified based on their variable inclusion proportions, defined as the proportion of times a variable was used to split a decision node across all trees. The MCMC process included a burn-in period of 10,000 iterations, followed by posterior sampling with every 20th draw retained (thinning), yielding 10,000 posterior samples. Convergence was evaluated using trace plots and autocorrelation functions, confirming adequate mixing and stability of posterior draws.

## Notes

### Competing Interest Statement

The authors have declared no competing interest.

### Clinical Trial

NCT03332537

### Clinical Protocols

https://pmc.ncbi.nlm.nih.gov/articles/pmid/29388674/

### Author Declarations

This study has been approved by the University of Connecticut-Storrs Institutional Review Board (IRB # H16-152; approval date: 9 September 2016) and conducted in accordance with the ethical standards outlined in the 1964 Declaration of Helsinki and its subsequent amendments. Informed consent was obtained from all subjects involved in the parent RCT study.

